# LONG-LASTING INSECTICIDE NETS OWNERSHIP AND MALARIA MORBIDITY IN KRACHI EAST MUNICIPALITY, GHANA

**DOI:** 10.1101/2022.05.18.22275276

**Authors:** Israel Wuresah, Siman Elmi, Martin Adjuiky

**Affiliations:** Department of Epidemiology and Biostatistics, School of Public Health University of Health and Allied Sciences, Ghana; Faculty of Health, B.Sc., Spec. Hons. Global Health (Policy, Management, and System), York University, Canada

**Keywords:** Malaria morbidity, LLINs ownership, LLINs usage

## Abstract

**Background:** Malaria-related morbidity and mortality are issues of great concern to public health globally though, a higher proportion of cases reside within Sub-Saharan Africa. The situation in Ghana though not new, is very disturbing, as millions of people especially children and pregnant women suffer severely from malaria. Seasonal chemoprevention and indoor residual spraying are among many measures deployed in the northern parts of Ghana with nationwide outreach and point distribution of LLINs across the country but reports from OPDs indicate millions of malaria cases annually.

**Objective:** To identify the levels of ownership and usage of the treated bed nets, and describe the relationship between ownership of LLINs and malaria morbidity.

**Methods:** The 30-cluster sampling method was deployed. Using both a modified WHO EPI survey method for more rural areas and a random walk sampling for more urban areas, each community had a listed starting point where the use of a spun pen determined the direction to conduct the surveys within the specified cluster. Selected households’ heads/representatives (any adult aged 18 years and above, in a household where the head is absent) participated voluntarily. STATA version 16.0 was used to run the statistical analysis and the results were presented in tables and figures.

**Results:** Findings revealed high levels of ownership of LLINs (73.4%) but moderately low usage levels (49.5%). Some other uses of LLINs (22.9%) aside from sleeping under them were identified. Malaria morbidity (59.6%) was also determined. Multivariate analysis results revealed statistically significant association between some socio-demographic characteristics and LLINs ownership including female sex (AOR = 2.1 (95% CI: 1.15, 3.87) p=0.016), being married (AOR = 3.4 (95% CI: 1.76, 6.74) p<0.001), cohabiting (AOR = 6.1 (95% CI: 2.15, 17.02) p=0.001) and being separated or divorced (AOR = 9.4 (95% CI: 1.09, 81.27) p=0.041). A positive correlation was identified between ownership of LLINs and their usage.

**Conclusion:** Despite high levels of ownership of LLINs, usage is minimal with a consequential effect on malaria morbidity. The study recommended service points and periodic household and/or community sensitization on LLINs usage as measures to increase usage levels.

## BACKGROUND

Malaria remains one of the major causes of morbidity and mortality globally. About 50% of the world’s population lives in malaria-endemic areas (Dayanand et al., 2017). Despite the many efforts put into controlling this infectious disease, the decline rate is still worrisome among some countries globally. The global mortality stood at 548,000 between 2012 and 2013 (Jemal & Ketema, 2019). The World Health Organization reported a significant decline in malaria cases over the years in the western world (World Health Organization (WHO), 2019) but the same cannot be said for Asia, Africa, and some countries in Latin America. About 500,000 cases and 50,000 malaria-related deaths were recorded in Pakistan in 2012 (Khattak, *et al*., 2013) and 1.5 million annual malaria cases were detected in India (Dayanand *et al*., 2017). Africa is noted for most malaria-related deaths globally, that is, 400,000 deaths of malaria, representing 90% of the world’s malaria burden (Mfuen *et al*., 2018; Dayanand *et al*., 2017). Castellanos *et al*., (2016) reported that there were 270,753 new cases of malaria in the mining communities of Colombia, representing 89.3% of the national malaria incidence between 2010 and 2013.

The reports of Ghana’s Demographic and Health Survey, GDHS (2014), revealed that the prevalence of malaria among children living in rural areas in Ghana stood at 38% as compared to the urban of 14%. Volta region recorded malaria prevalence among children to be 36.6% according to the same reports.

Instigating the need for this research is the continued influx of malaria cases recorded at outpatient departments (OPDs) in Ghanaian health facilities. Deku *et al*., (2018) reported 2.3 million cases of malaria at the OPDs. Children aged below 5 years and pregnant women are the most affected groups (Lamptey *et al*., 2018). Malaria in Ghana contributes to 17.6% of the total OPD attendance, 13.7% of total admissions, and 3.4% of maternal deaths (Anabire *et al*., 2019). Meanwhile, the distribution of LLINs has become a routine in the country.

The non-inverse association, however, could be due to many extraneous factors. This study determined the relationship between ownership of LLINs and malaria morbidity by investigating coverage (ownership), levels of usage, and malaria prevalence in the municipality.

## METHODS

### Study design

A household-based cross-sectional survey was conducted to examine the relationship between LLINs ownership and malaria prevention in Krachi-East Municipality. The study gathered data from the settings of participants through the use of a pretested structured questionnaire. The questionnaire was designed to elicit information on socio-demographic characteristics, household characteristics, as well as ownership and usage of LLINs. The 30-cluster sampling technique was used due to the large size of the municipality as a means of ensuring that the data was collected from the varied population attributes within the municipality.

### Study population and procedure

This study included adults aged 18 years and above who have consented to participate in the study by signing or thumb printing the consent form within the chosen households of the selected communities in the municipality.

Through the use of an interview, an adjusted questionnaire from Ghana’s Malaria Indicator Survey (MIS) was administered first to a similar population in a different region as a pretesting phase and then administered to the Krachi-East Municipality afterwards.

### Inclusion and Exclusion Criteria Inclusion Criteria

The study population consisted of both males and females 18 years of age and above, who resided within the Krachi-East Municipality. Those residing within the municipality lived in one residence before and throughout the study. Lastly, said participants were able to speak for themselves as well as consent or assent to participate in the study.

### Exclusion Criteria

The study population did not consist of males and females below the age of 18. Those who do not reside and or reside in multiple residences in the Krachi-East Municipality were also excluded, as well as individuals that were not able to give their consent for participation were also excluded from the study.

### Ethical considerations

Ethical approval was obtained from the University of Health and Allied Sciences Review Ethics Committee (UHAS-REC). Due processes were followed before, during, and after the study.

### Statistical Analysis

Responses gathered in the KoboCollect software were extracted into an excel sheet and exported to STATA^®^ version 16.0 for analysis after cleaning. For descriptive purposes, Microsoft Excel was used to generate charts for better understanding. Categorical variables were summarized into percentages and cross-tabulations and continuous variables were summarized into means and standard deviations. A chi-square analysis was run to determine the relationship between ownership of LLINs, levels of usage, and malaria prevalence. Binary logistic regression was performed to get the odds of LLINs ownership to determine the strength of the association where p-values of less than 0.05 at a 95% confidence interval were deemed statistically significant.

## RESULTS

A total of 297 respondents sampled from 28 communities were enrolled in the study. Ninety-eight (88) respondents (29.6%) were aged 18-29 years, 65 (22.2%) aged 30-39, 66 respondents were aged 40-49 (22.2%) and 77 respondents (25.9%) were aged 50 years and above. The mean age of respondents was 40.5 (15.4).

Female respondents dominated the study with 191 (64.3%) of 297 respondents as compared to the male respondents, 106 (35.7%).

Averagely, each household had a population of 7.1 (4.3) Household sizes ranged from ≤ 2 (20, 6.7%), 3-5 (103, 34.7%), 6-8 (92, 31.0%) and ≥ 9 (82, 27.6%).

One hundred and thirty-six (136) households (45.8%) used ≤ 2 rooms for sleeping, 105 (35.4%) used 3-5 rooms, and 56 (18.9%) used ≤ 5 rooms for sleeping. On average, 3.20 (2.46) sleeping-in rooms were recorded.

### Ownership and usage of LLINs, and Malaria Prevalence

Of the 297 households, 218 (73.4%) had at least one LLIN and 79 (26.6%) were without LLINs (Table 2.0). Averagely, each household has a 2.3 (2.3) number of nets.

**Table 1.0:** Sociodemographic characteristics of respondents (N=297)

| **Variable** | **Frequency** | **Percentage** |
| --- | --- | --- |
| **Age (years) (S.D)** | 40.5 (15.4) |  |
| 18-29 | 88 | 29.7 |
| 30-39 | 66 | 22.2 |
| 40-49 | 66 | 22.2 |
| 50 and above | 77 | 25.9 |
| **Sex** |  |  |
| Male | 106 | 35.7 |
| Female | 191 | 64.3 |
| **Ethnicity** |  |  |
| Ewe | 79 | 26.6 |
| Konkomba/Basare | 51 | 17.2 |
| Guan | 49 | 16.5 |
| Akan | 26 | 8.8 |
| Ga/Adanbge | 21 | 7.1 |
| Others | 71 | 23.9 |
| **Religion** |  |  |
| Christian | 226 | 76.1 |
| Islamic | 30 | 10.1 |
| Traditional | 28 | 9.4 |
| Others | 13 | 4.4 |
| **Occupation** |  |  |
| No occupation | 44 | 14.8 |
| Social worker | 16 | 5.4 |
| Farmer | 120 | 40.4 |
| Trader | 73 | 24.6 |
| Others | 44 | 14.8 |
| **Marital status** |  |  |
| Never married | 58 | 19.5 |
| Married | 190 | 64.0 |
| Cohabiting | 38 | 12.8 |
| Divorced/Separated/Widowed | 11 | 3.7 |
| **Educational level** |  |  |
| No formal education | 106 | 35.7 |
| Basic school | 57 | 19.2 |
| Junior High School | 79 | 26.6 |
| Senior High School | 42 | 14.1 |
| Tertiary | 13 | 4.4 |
| **Income (monthly)(GHC)** |  |  |
| No income | 51 | 17.2 |
| Less 100 | 90 | 30.3 |
| 100 –499 | 120 | 40.4 |
| 500 –999 | 25 | 8.4 |
| 1000 or more | 11 | 3.7 |
| **Community type** |  |  |
| Rural | 138 | 46.5 |
| Urban | 159 | 53.5 |

**Table 2.0:**
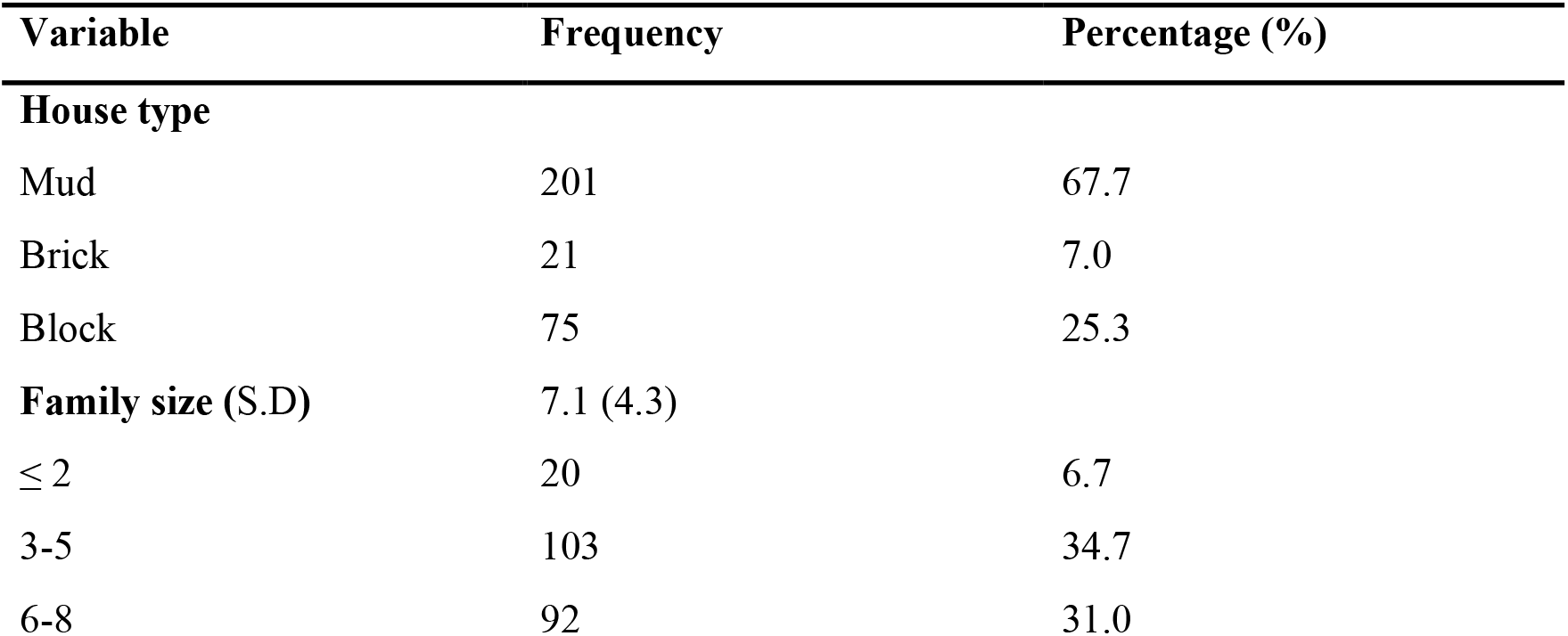
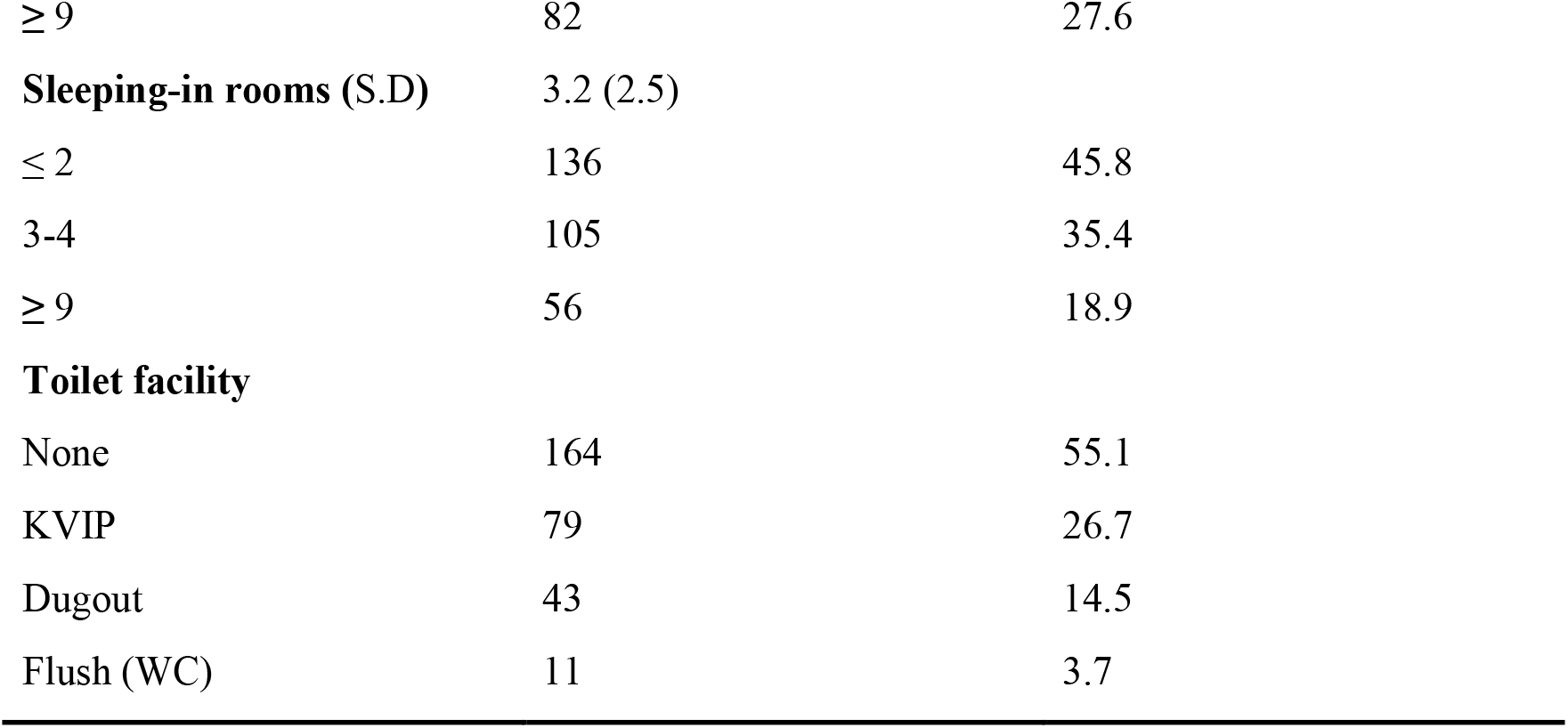
Household Characteristics of respondents (N=297)

**Table 3.0:** Ownership and Usage of LLINs, and Malaria prevalence.

| variable | Frequency N=297 | Percentage % |
| --- | --- | --- |
| **LLINs ownership** |  |  |
| No | 79 | 26.6 |
| Yes | 218 | 73.4 |
| **LLINs usage** |  |  |
| No | 150 | 50.5 |
| Yes | 147 | 49.5 |
| **Other uses of LLINs (n=68)** |  |  |
| Gardening | 33 | 48.5 |
| Fishing | 19 | 27.9 |
| Others | 16 | 23.5 |
| **Malaria morbidity** |  |  |
| No | 120 | 40.4 |
| Yes | 177 | 59.6 |

**Table 4.0:** Bivariate and Multivariate analysis of socio-demographic characteristics of respondents, LLINs ownership, and Utilization.

| Variable | LLINs Ownership | | $\chi^2$ | p-value | COR [95% CI], p-value | AOR (95% CI), p-value |
| --- | --- | --- | --- | --- | --- | --- |
|  | Yes | No |  |  |  |  |
|  | n (%) | n (%) |  |  |  |  |
| **Age** |  |  |  |  |  |  |
| 18-29 | 60 (68.2) | 30 (30.6) | 2.9.7 | 0.412 |  |  |
| 30-39 | 52 (78.8) | 14 (21.2) |  |  |  |  |
| 40-49 | 51 (77.3) | 15 (22.7) |  |  |  |  |
| 50 and above | 55 (71.4) | 22 (28.6) |  |  |  |  |
| **Sex** |  |  |  |  |  |  |
| Male | 68 (64.2) | 38 (35.8) | 7.2 | **0.007\*** | 1 | 1 |
| Female | 150 (78.5) | 41 (21.5) |  |  | 2.0 [1.21 – 3.46], **0.008** | 2.1[1.15 – 3.87], **0.016** |
| **Ethnicity** |  |  |  |  |  |  |
| Ewe | 63 (79.8) | 16 (20.2) | 4.4 | 0.490 |  |  |
| Konkomba/Basare | 39 (76.5) | 12 (23.5) |  |  |  |  |
| Guan | 32 (65.3) | 17 (34.7) |  |  |  |  |
| Akan | 20 (76.9) | 6 (23.1) |  |  |  |  |
| Ga/Dangbe | 15 (71.4) | 6 (28.6) |  |  |  |  |
| Others | 49 (69.0) | 22 (31.0) |  |  |  |  |
| **Religion** |  |  |  |  |  |  |
| Christianity | 173 (76.6) | 53 (23.4) | 7.2 | 0.065 |  |  |
| Islamic | 20 (66.7) | 10 (33.3) |  |  |  |  |
| Traditionalist | 19 (67.9) | 9 (32.1) |  |  |  |  |
| Others | 6 (46.1) | 7 (53.9) |  |  |  |  |
| ***Marital Status*** |  |  |  |  |  |  |
| Single | 30 (51.7) | 28 (48.3) | 18.5 | < **0.001\*** | 1 | 1 |
| Married | 147 (77.4) | 43 (22.6) |  |  | 3.2 [1.72 – 5.91], <**0.001** | 3.4 [1.76 – 6.74], <**0.001** |
| Co-habiting | 31 (81.6) | 7 (18.4) |  |  | 4.1 [1.57 – 10.90], **0.004** | 6.0 [2.15 – 17.02], **0.001** |
| Divorced/Separated | 10 (90.9) | 1 (9.1) |  |  | 9.3 [1.12 – 77.69], **0.039** | 9.4 [1.09 – 81.27], **0.041** |
| ***Educational Level*** |  |  |  |  |  |  |
| No formal education | 83 (78.3) | 23 (21.7) | 5.5 | 0.237 |  |  |
| Basic school | 42 (73.7) | 15 (26.3) |  |  |  |  |
| JHS | 58 (73.4) | 21 (26.6) |  |  |  |  |
| SHS | 25 (59.5) | 17 (40.5) |  |  |  |  |
| Tertiary | 10 (76.9) | 3 (23.1) |  |  |  |  |
| ***Occupation*** |  |  |  |  |  |  |
| No occupation | 31 (70.4) | 13 (29.6) | 11.3 | **0.024\*** | 1 | 1 |
| Social worker | 15 (93.8) | 1 (6.2) |  |  | 6.3 [0.75 – 52.68], 0.090 | 7.8 [0.88 – 69.75], 0.065 |
| Farmer | 86 (71.7) | 34 (28.3) |  |  | 1.1 [0.50 – 2.27], 0.879 | 1.0 [0.41 – 2.39], 0.987 |
| Trader | 60 (82.2) | 13 (17.8) |  |  | 1.9 [0.80 – 4.68], 0.143 | 1.7 [0.67 – 4.43], 0.257 |
| Others | 26 (59.1) | 18 (40.9) |  |  | 0.6 [0.25 – 1.46], 0.27 | 0.6 [0.24 – 1.68], 0.355 |
| ***Income*** |  |  |  |  |  |  |
| None | 36 (70.6) | 15 (29.4) | 7.7 | 0.103 |  |  |

| < 100 | 65 (72.2) | 25 (27.8) |  |  |  |  |
| --- | --- | --- | --- | --- | --- | --- |
| 100 – 499 | 84 (70.0) | 36 (30.0) |  |  |  |  |
| 500 – 999 | 22 (88.0) | 3 (12.0) |  |  |  |  |
| >= 1000 | 11 (100.0) | 0 (0.0) |  |  |  |  |
| *Community* |  |  |  |  |  |  |
| Rural | 108 (78.3) | 30 (21.7) | 3.1 | 0.077 |  |  |
| Urban | 110 (69.2) | 49 (30.8) |  |  |  |  |
| Variable | LLINs Utilization | | $\chi^2$ | p-value | COR (95% CI), p-value | AOR (95% CI), p-value |
|  | Yes | No |  |  |  |  |
|  | n (%) | n (%) |  |  |  |  |
| *Community* |  |  |  |  |  |  |
| Rural | 54 (39.1) | 84 (60.9) | 11.0775 | **0.001\*** | 1 | 1 |
| Urban | 93 (58.5) | 66 (41.5) |  |  | 2.2 [1.38 – 3.49], **0.001** | 2.8 [1.62 – 4.73], <**0.001** |
| *Ownership of LLINs* |  |  |  |  |  |  |
| No | 20 (25.3) | 59 (74.7) | 25.1704 | <**0.001\*** | 1 | 1 |
| Yes | 127 (58.3) | 91 (41.7) |  |  | 4.1 [2.32 – 7.31], < **0.001** | 4.7 [2.49 – 8.90], <**0.001** |

Usage of the LLNIs was recorded among 147 households (49.5%) and non-usage responses of 150 (50.5%) were recorded also (Table 2.0). Other uses for the LLINs were either observed or reported in 68 (22.9%) households. Of the 68 respondents, 33 (48.5%) used the LLINs for gardening, 19 (27.9%) used the nets for fishing, and 16 (23.5%) used them for hen coops, and drying food commodities among others. Presumptuously, 147 households (49.5%) used the LLINs among 218 (73.4%) who owned an LLIN.

One hundred and twenty (120, 40.4%) respondents had no history of malaria in the past 12 months before the study but 177 (59.6%) reported having had malaria in the said period.

**Figure 1:**
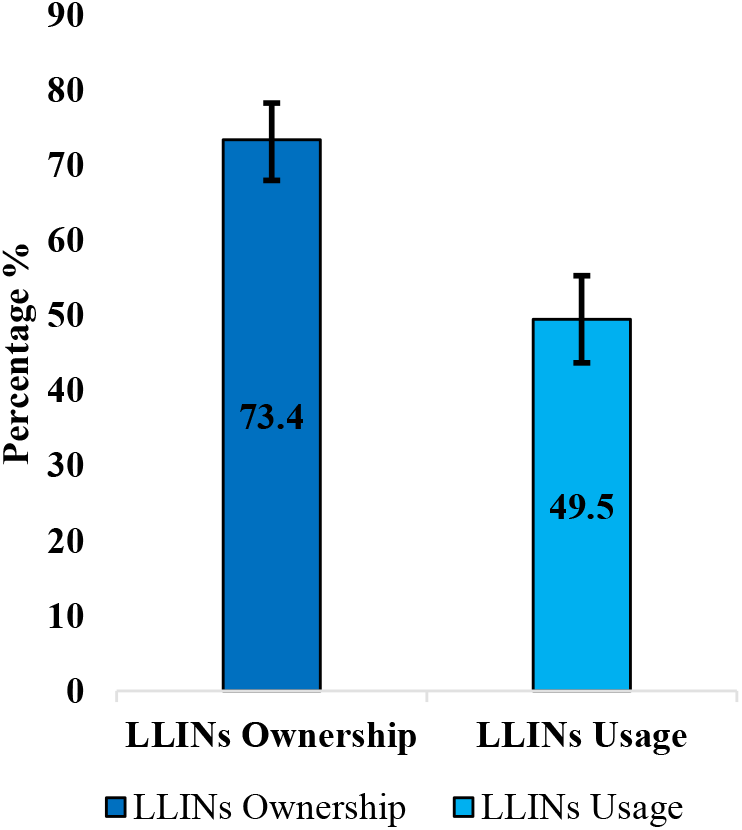
Ownership and Usage of LLINs.

**Figure 2:**
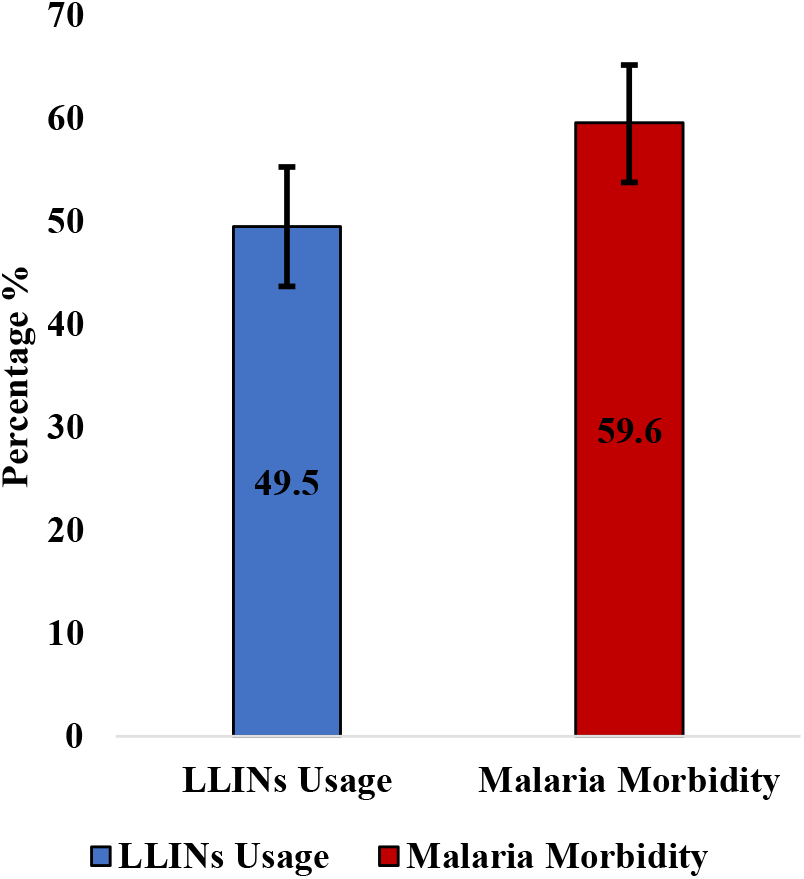
LLINs Usage and Malaria prevalence.

### Bivariate and Multivariate analysis of socio-demographic characteristics of respondents, LLINs ownership, and Utilization

The indicators of LLINs ownership showed no association with the various age groups, ethnicities, various religious groups, all levels of education, income levels, and community types. However, there was a significant association between being a female (AOR=2.1 (95% C.I: 1.15 – 3.87) p=0.016) and owning LLINs. Married participants (AOR=3.4 (95% C.I: 1.76 – 6.74) p=<0.001), participants cohabiting (AOR=6.1 (95% C.I: 2.15 – 17.02) p=0.001) and divorced/separated participants (AOR=9.4 (95% C.I: 1.09 – 81.27) p=0.041) were more likely to own a LLINs. It was likely for social workers, farmers, traders, and other occupations to own LLINs since there was an association between these covariates and ownership of LLINs at the bivariate levels. Association at the multivariate level was however insignificant.

Utilization of LLINs was highly possible (AOR=2.8 (95% C.I: 1.62 – 4.73) p=<0.001) among participants in urban Krachi-East. There was also a significant association (AOR=4.7 (95% C.I: 2.49 – 8.90) p=<0.001) between owning an LLIN and using it.

## DISCUSSION

### Ownership of Long-lasting insecticidal nets (LLINs)

Globally, long-lasting insecticidal nets have contributed to the reduction of malaria burdens. So far, they are the most cost-effective and a core strategy used widely in malaria-endemic and epidemic-prone areas. The study discovered that all LLINs owned by households interviewed were obtained through mass distribution campaigns championed by the US President’s Initiative. This finding correlates with the Global Technical Strategy for malaria as envisaged by the WHO, to ensure universal coverage of LLINs for all people at risk of malaria (WHO, 2017) by obtaining and sustaining global coverage for malaria control with long-lasting insecticidal nets through mass free net distribution campaigns and sustained distribution through multiple channels.

Several studies purported that the educated populace owned LLINs more (Ernst *et al*., 2016; Axame *et al*., 2016). Contrary to their findings, LLINs ownership was reported more among people with no formal education, females, farmers, married respondents, and respondents who resided in urban Krachi-East. The implication of these findings may be that a person’s level of education is unlikely to influence their ownership of a treated bed net.

### Usage of long-lasting insecticidal net

Results from the study showed that net utilization was low compared to the proportion of ownership. Findings from the study are that religion, marital status, and locality type (be it urban or rural) impact LLINs usage. The findings agree with Gryseels *et al*., (2019), which projected that usage of LLINs could be affected by some sociocultural, educational, economic, and environmental factors. Usage of LLINs is poor as identified by this study, despite high ownership among residents of the municipality. Several other studies discovered similar findings (Mukhopadhyay *et al*., 2016; Khanam *et al*., 2018; Anuse, Sahu, Subramanian & Gunasekaran, 2015; Raaghavendra, 2017; Painstil *et al*., 2014).

### Malaria morbidity

Undoubtedly, some progress has been made in the prevention and control of malaria. Feachem *et al*., (2019), stated that more than half of the countries in the world are free from malaria. However, the cases are still overwhelming comparing the efforts made so far. Year in and year out, campaigns are being launched, and interventions are being rolled out, yet cases of malaria keep falling and rising; this is indicative of no progress being made. We discovered that out of every ten (10) persons interviewed, six (6) had had malaria before the study. Snow et al., (2017) observed inconsistent and meagre decline rates of malaria across the belts of high malaria transmission areas in Africa from 1990 to 2015. In furtherance of these findings is Africa accounting for about 93% of malaria cases globally and nearly 85% of global malaria fatality, (WHO, 2019), despite the consistent deployment of LLINs. The rhetorical question remains, do malaria prevention and control require a lot more distribution of LLINs, or does it require behaviour change interventions or both?

## Conclusion

Although a lot of people believe LLINs are effective for controlling malaria, usage is still low. The maximum use of LLINs is integral to achieving an all-out control and elimination of malaria using LLINs. It is therefore concluded that low usage of LLINs is an associated factor with the high level of malaria morbidity in the Krachi East municipality.

### Recommendations

Using the previously collected data what can be acknowledged is that the ownership of LLINs has shown promising results in eradicating malaria within Ghana. Though due to the overall lack of usage and or misuse of LLINs there is still a gap in the quality of life for individuals within this region. This gap must be filled through service points and periodic household and/or community education, periodic cues presentations via mass media, and other viable media to raise awareness of good practices about LLINs. Along with such, further measures such as indoor residual spraying, and community fumigation exercises should be taken to complement LLINs to make up for low usage of the LLINs, particularly among populations who find it difficult to sleep under the net. Future research may explore the factors accounting for the low usage of LLINs in the municipality.

## Data Availability

The datasets generated during and/or analyzed during the current study are available from the corresponding author on reasonable request.

https://kf.kobotoolbox.org/#/forms/amTCqjMAg7MUHSFSiQNaeW

